# Mortality in Eastern Democratic Republic of the Congo: A Population-Based Survey Following the 2025 M23 Offensive and Humanitarian Funding Withdrawal

**DOI:** 10.64898/2026.07.10.26357778

**Authors:** Jennifer OKeeffe, Augustin Gang Karume, Les Roberts

**Author notes:** **Transparency Declaration** The lead author^*^ affirms that this manuscript is an honest, accurate, and transparent account of the study being reported; that no important aspects of the study have been omitted; and that any discrepancies from the study as planned (and, if relevant, registered) have been explained. **Ethical Considerations** The protocol was approved by the Catholic University of Bukavu (Approval: UCB/CIES/PB/013/2025). We obtained administrative authorization at the provincial, zonal, and area levels authorities controlling that area. Due to low literacy and security constraints, we conducted verbal informed consent. Data were anonymized and stored on encrypted, password-protected servers. **Data Sharing** Due to the sensitive nature of the study, data are not publicly shared. Requests for data may be submitted to the corresponding author and considered on a case-by-case basis. **Patient and Public Involvement** Due to the sensitive nature of the study, patients and the public were not involved in design or conduct of the study due to potential personal risk. **Competing Interests** All authors have no competing interest to declare.

## Abstract

**Introduction:** In early 2025, North and South Kivu Provinces in the Democratic Republic of the Congo (DRC) underwent a major geopolitical shift marked by the concurrent abrupt withdrawal of humanitarian support and large-scale military takeover by the M23 armed group. This study sought to quantify mortality before and after the humanitarian and security collapse.

**Methods:** We conducted a retrospective, two-stage cluster household survey comparing mortality in the pre- and post-crisis periods. Difference-in-differences were analyzed using survey-weighted Poisson regression with a log-link and person-time offset to estimate adjusted incident rate ratios for crude, under-five, and sex-specific mortality. We interviewed community leaders and health facility staff as key informants to provide qualitative data.

**Results:** The adjusted risk of death was 2.02 times higher in the post-crisis than the pre-crisis period (95% CI 1.07, 3.80; p=0.03). Crude mortality rose from a pre-crisis adjusted marginal mean of 0.38 (95% CI 0.22, 0.53) to 0.74 (95% CI 0.56, 0.92) deaths/10,000/day. The increase in mortality was consistent across DRC-Government and M23 controlled areas. Results indicate that 165,391 (95%CI: 63,557, 267,225) excess deaths occurred in the post-crisis period, equating to 245,397 annual excess deaths.

**Conclusion:** The crisis in DRC is driven by the convergence of humanitarian funding withdrawals and escalating conflict. Averting further preventable deaths requires large-scale restoration of humanitarian assistance. The resources needed to do so represent a small fraction of global economic capacity.

**What is already known on this topic:** Elevated population mortality has been widely documented in periods of conflict. Eastern DRC is currently experiencing an escalation of conflict and simultaneous withdrawal of humanitarian aid, indicating increased risk of preventable death.

**What this study adds:** This study will provide the first published mortality estimates based on primary data collection for eastern DRC since the fall of Goma in January 2025.

**How this study might affect research, practice or policy:** This study measured a doubling of population mortality over a 7-month period, meeting the emergency threshold of a humanitarian crisis. Declaration of a crisis should trigger urgent response from humanitarian actors to avert further preventable death.

## Introduction

The Democratic Republic of Congo (DRC) ranks 171 out of 193 countries on the 2025 Human Development Index with a life expectancy of 61.9 years^1^. An estimated 85% of the population lives below the poverty line of 3 USD per day. When measured by total population affected, the DRC represents the world’s largest food insecurity crisis^2^.

Competition over geological wealth has resulted in decades of armed conflict, particularly in the mineral-rich eastern provinces of North and South Kivu. The second Congolese war, from 1998-2003, resulted in an estimated 5.4 million excess deaths, the deadliest conflict since WWII.^3^ At the end of 2024, roughly 7.8 million people were internally displaced, with an additional 2.9 million returnees.^4^

Chinese firms control up to 80% of primary mining concessions^5^ while the United States has established strategic agreements to secure supply chains.^6^ Neighboring Rwanda and Uganda have active troops in the DRC, citing national security concerns of non-state armed groups. However, United Nations (UN) experts report their presence primarily serves as a means to export hundreds of millions of dollars of mineral resources per year^7^

In 2013, the Rwandan-backed March 23 Movement (M23) seized Congolese territory before withdrawing when US-led sanctions were imposed against Rwandan Government officials.^8^ In 2025, the DRC underwent a major geopolitical shift marked by the abrupt withdrawal of humanitarian support followed by renewed large-scale military incursions by M23. In a reversal of decades of foreign policy, the United States Agency for International Development (USAID), enacted drastic funding cuts, including immediate stop-work orders which froze life-saving assistance^9^. U.S. share of humanitarian funding in the DRC shrunk from 68% to under 20%^10^.

Following the reduction in financial aid, the M23, with its political arm, Alliance Fleuve Congo (AFC), launched an offensive that quickly overwhelmed the Congolese Armed Forces (FARDC), capturing unprecedented territory, and cutting off key supply routes and humanitarian corridors. Unable to contain the advance, the Congolese government integrated the Wazalendo, a loose coalition of armed groups, as official military reserves.^11^ All parties to the conflict have committed widespread human rights abuses, including summary executions, torture, sexual violence, disappearances, recruitment of child soldiers, extortion, and forced displacements.^12, 13^

The environment of violence and deterioration of the health and humanitarian systems has rendered increased mortality and morbidity inevitable. While field reports and rapid assessments suggest a substantial rise in preventable deaths^14^, reliable epidemiological data are limited due to the insecurity and withdrawal of humanitarian actors^10^.

This study sought to quantify mortality before and after the security and humanitarian collapse in North and South Kivu, aiming to document the human consequences of the simultaneous conflict escalation and funding retraction in Eastern DRC.

## Methods

### Survey Design

We conducted a retrospective cross-sectional household survey in North and South Kivu provinces,eastern DRC using a two-stage cluster design. Data collection took place from August 1 to September 15, 2025. The analysis compared mortality across two periods: 1) a pre-crisis period from August 1, 2024, to January 31, 2025, and a post crisis period from February 1, 2025. the date of interview, between July 31 to Sept 29. Local event calendars were used to anchor recall dates.

### Sample Size

The sample size was powered to independently detect a doubling of mortality between the pre- and post-crisis periods in each province. Given that the official UN estimate of 9 deaths per 1000 population per year^15^ is likely an underestimate, we assumed a pre-crisis rate of 18 deaths per 1000 per year. With 95% significance, 80% power, and ±60% relative precision for the baseline, the minimum required sample was 1,088 individuals per province. This was doubled to account for clustering, household non-response, and cluster inaccessibility. Assuming an average household size of five and 15 households per cluster, a stratified design of 35 clusters per province (approximately 2,625 people per province) was determined to be sufficient.

### Sampling Strategy

In the first sampling stage, 35 clusters were selected probability proportional to size for each province based on 2025 Ministry of Health population estimates. In the second stage, larger villages or neighborhoods were systematically segmented until an area of 40 to 60 households was obtained. Villages estimated to be under 75 households were sampled without segmentation. All households in the sample area were enumerated, and teams randomly selected the starting household. They surveyed the 14 subsequent households to the left without replacement. The sampling approach was chosen to minimize survey team exposure in insecure areas. During cluster selection, potential logistic and security constraints were not considered, and inaccessible clusters were not replaced. In the final selection, clusters were approximately equally distributed between DRC government and M23-controlled areas.

### Survey Population

All individuals residing in the selected households during the recall period, including internally displaced persons (IDPs), were eligible. A household was defined as a group sharing a common head of household and eating from the same pot for the majority of the past 30 days. All in- and out migrations to the household lasting more than two weeks were recorded. In polygamous households, adult men were assigned to the household where they ate most of their meals in the past 30 days, If meals were reportedly shared equally they were assigned to the household of the first wife.

### Inclusion/Exclusion Criteria

To be considered eligible for the study, households needed a representative 18+ years (or if serving as head of household, 15+ years) who was present at the time of interview and could provide information over the entire recall period. Households were excluded if they refused participation, lacked an eligible representative, or remained absent during data collection. Teams revisited absent households on subsequent days in accessible areas or later the same day in hard-to-reach clusters.

### Data collection and entry

We collected household data on demographics, vital events, displacement, and humanitarian assistance, measuring nutritional status in children 6–59 months via mid-upper arm circumference. Interviews concluded with an open-ended question asking if the participant had anything else to share. Where feasible, we conducted key informant interviews with health facility staff on service utilization, supply chains, and financial aid, and with community leaders on conflict and displacement. The tool was developed in French and administered in Swahili or Mashi. Data were double entered into a Kobo database from paper forms. Enumerators completed three days of training on study ethics, procedures, and psychological first aid, followed by a one-day pilot.

### Statistical Analysis

Data were analyzed in R (version 4.2.2) using survey-weighted Poisson regression with a log link and person-time offset. Missing data was less than 0.1% of We employed a difference-in-differences (DiD) approach to estimate adjusted incident rate ratios (aIRR) for mortality, selecting the Poisson distribution based on Akaike Information Criterion (AIC) comparisons (Table S1). Missing data comprised <0.01% of key variables, which were removed from analysis.

The final model adjusted for province (North or South Kivu) and accessibility (a binary indicator with hard-to-reach clusters defined as needing any of: >1 attempt to reach, >1 day of travel, local security escorts, or additional negotiation or authorization from officials or non-state armed groups). The model additionally included main effects for period (pre- vs post-crisis) and territorial control (DRC-Gov vs M23-AFC) with an interaction term for the DiD effect. We conducted sensitivity analyses to assess model specification, covariate influence, cluster outliers (exclusion of highest mortality cluster), and temporal measurement bias (washout analysis excluding the months prior and following the crisis cut-off date) (Table S4).

While the aIRR serves as the primary effect measure, we present adjusted mortality rates as a descriptive measure to aid interpretation of results within a broader global and historical context. Crude, under-five and sex-disaggregated rates are reported as deaths per 10,000 per day, with person-time adjusted for births, deaths, and migration.

Excess deaths were calculated by subtracting the expected deaths, based on the pre-crisis marginal mean rate, from the observed marginal mean rate during the post-crisis period. Standard errors were estimated using the delta method to derive 95% confidence intervals. We also present descriptive statistics on population structure (Figure S1), birth rates (Table S5), and proximate determinants of mortality, morbidity and service utilization (Table S6).

## Results

### Study Population and Baseline Characteristics

Between July 31 and September 26, 2025, we surveyed 899 households across 60 clusters in North and South Kivu, with 93.3% response rate (Table 1.). DRC-Gov and M23-AFC controlled areas had statistically similar samples by number of households surveyed, total population, and person-time. The total ever-present study population across the recall period included 6,736 individuals, with 2,230,502 person-days at risk (mean of 331.1 days). Demographics were also similar across strata, with a median age of 15 years (IQR 7–28), 52.1% (n=3,488) female, and 16.2% (n=1,072) children under five.

**Table 1.** Survey, household, and demographic characteristics of the study population by territorial control North and South Kivu, DRC, August 2024 – August 2025.

|  | Total | DRC – Gov | M23 – AFC |
| --- | --- | --- | --- |
| **Survey Characteristics** |  |  |  |
| Clusters sampled | 60 | 26 | 34 |
| Number of households surveyed | 899 | 389 | 510 |
| % household non-response (n) | 6.8 (61) | 8.2 (32) | 5.7 (29) |
| Mean household size (95%CI) | 6.9 (6.7, 7.1) | 6.6 (6.3, 6.9) | 7.1 (6.8, 7.4) |
| **Population Sample Size** |  |  |  |
| Population at mid-point | 5,702 | 2,340 | 3,362 |
| Total person-days at risk | 2,230,502 | 933,674 | 1,296,828 |
| Average days at risk per person | 331.1 | 351.5 | 317.9 |
| **Demographics** |  |  |  |
| Age, Median (IQR) | 15 (7,28) | 15 (7, 30) | 15 (7, 27) |
| Under five years, n (%) | 1,072 (16.2) | 404 (15.3) | 668 (16.7) |
| Female, n (%) | 3,488 (52.1) | 1,344 (50.7) | 2,144 (53.1) |
| **Vital Events** |  |  |  |
| Closed pregnancies (n) | 265 | 109 | 156 |
| Live births n (%) | 208 (78.5) | 73 (67.0) | 135 (86.5) |
| All-age Deaths (n) | 128 | 62 | 66 |
| Under-five Deaths, n (%) | 39 (31.0) | 12 (19.4) | 27 (40.9) |
| Female Deaths, n (%) | 49 (38.9) | 23 (38.3) | 26 (39.4) |
| **Population Movement** |  |  |  |
| Total population (ever-present) | 6,736 | 2,656 | 4,080 |
| Arrivals, n (%) | 919 (13.6) | 278 (10.5) | 641 (15.7) |
| Departures, n (%) | 814 (12.1) | 221 (8.3) | 593 (14.5) |
| Displaced over recall period, n (%) | 1,056 (15.7) | 291 (11.0) | 765 (18.8) |

*Key Findings: The survey included 899 households across 60 clusters. The population was young with a median age of 15 (IQR 7, 28). Displacement was high, with 15.7% of the population displaced over the recall period.*

Over the recall period, we recorded 208 live births and 128 deaths, of which 31.0% (n=39) were children under five. Population instability was substantial across the region with 13.6% (n=919) arriving into a household and 12.1% (n=814) departing. Displacement was the primary reported reason for migration, affecting 15.7% (n=1,056) of the sample. It was more frequent in M23–AFC-controlled zones, where 18.8% (n=765) were displaced compared to 11.0% (n=291) in DRC Government-held areas (p<0.001).

### Mortality Difference-in-Differences Analysis

The risk of death was 2.02 times higher in the post-crisis than the pre-crisis period (95% CI 1.07, 3.80; p=0.03) after adjusting for province and accessibility (Table 2). The CMR rose from a pre-crisis adjusted marginal mean of 0.38 (95% CI 0.22, 0.53) to 0.74 (95% CI 0.56, 0.92) deaths/10,000/day (Figure 1). Female mortality risk increased 2.65 times (95% CI 1.15,6.12; p=0.02) post-crisis corresponding to a rate increase from 0.28 (95% CI 0.13, 0.43) to 0.57 (95% CI 0.34, 0.79) deaths/10,000/day. The crude unadjusted risk was consistent with adjusted risk at 2.04 (95%CI: 1.08, 3.83; p=0.03) times higher in the post-crisis period. Female unadjusted mortality risk was 2.68 (95%CI: 1.16, 6.18) times higher in the post-crisis period. Changes in under-five and male mortality were not statistically significant (U5MR: p=0.26; Male MR: p=0.15). We documented 11 persons who had disappeared, 6 of whom were children under 18 years. These are potential, but not confirmed, deaths, and were not counted in population mortality. Complete model results are shown in Table S2.

**Table 2.** Adjusted Pre- and Post-Crisis Incident Rate Ratios and Mortality Rates in North and South Kivu, DRC, August 2024-August 2025. *Key Findings: For crude mortality, the risk of death was 2.02 (95%CI: 1.07, 3.80; p = 0.03) higher in the post-crisis period. The change in risk was significant in females who faced an increase in risk of 2.65 (95%CI: 1.15, 6.12; p = 0.02) times compared to the baseline. Changes in under-five and male mortality did not reach significance (p>0.05.) Notably, the interaction terms for the post-crisis period x M23-controlled areas were non-significant across all groups (p > 0.05), indicating mortality trends were consistent across territories*

| | Post-Crisis Period<br>aIRR (95%CI) | Post-Crisis Period<br>aIRR p-value | Post-Crisis × M23<br>p-value | Pre-Crisis Mortality<br>Rate (95%CI) | Post-Crisis Mortality<br>Rate (95%CI) |
| --- | --- | --- | --- | --- | --- |
| Crude | 2.02 (1.07, 3.80) | 0.03 | 0.88 | 0.38 (0.22, 0.53) | 0.74 (0.56, 0.92) |
| Under-Five | 2.23 (0.55, 9.10) | 0.26 | 0.51 | 0.67 (0.15, 1.18) | 1.15 (0.58, 1.72) |
| Female | 2.65 (1.15, 6.12) | 0.02 | 0.36 | 0.28 (0.13, 0.43) | 0.57 (0.34, 0.79) |
| Male | 1.74 (0.82, 3.68) | 0.15 | 0.61 | 0.47 (0.25, 0.69) | 0.92 (0.70, 1.14) |

**95%CI:** = Confidence Interval; **aIRR**=Adjusted Incidence Rate Ratio; **DiD** = Difference- in-Differences
**Rate Units:** Adjusted mortality rates are expressed as deaths per 10,000 people per day.
**Statistical Analysis:** Estimates were derived from survey-weighted Poisson regression models adjusted for province and accessibility. The analysis incorporated inverse probability weights and cluster-robust standard errors to account for the sampling design and intra-cluster correlation at the village and household levels. The aIRR reflects the regional mortality change between pre-crisis (Aug 2024–Jan 2025) and post-crisis (Feb–Aug 2025) periods. The interaction p-value tests whether changes in mortality differed significantly between DRC Government and M23-AFC territorial control. Mortality rates represent adjusted marginal mean rates (deaths/10,000/day) from the survey-weighted Poisson models.
**Significance:** Statistical significance is set at $p<0.05$

**Figure 1.**
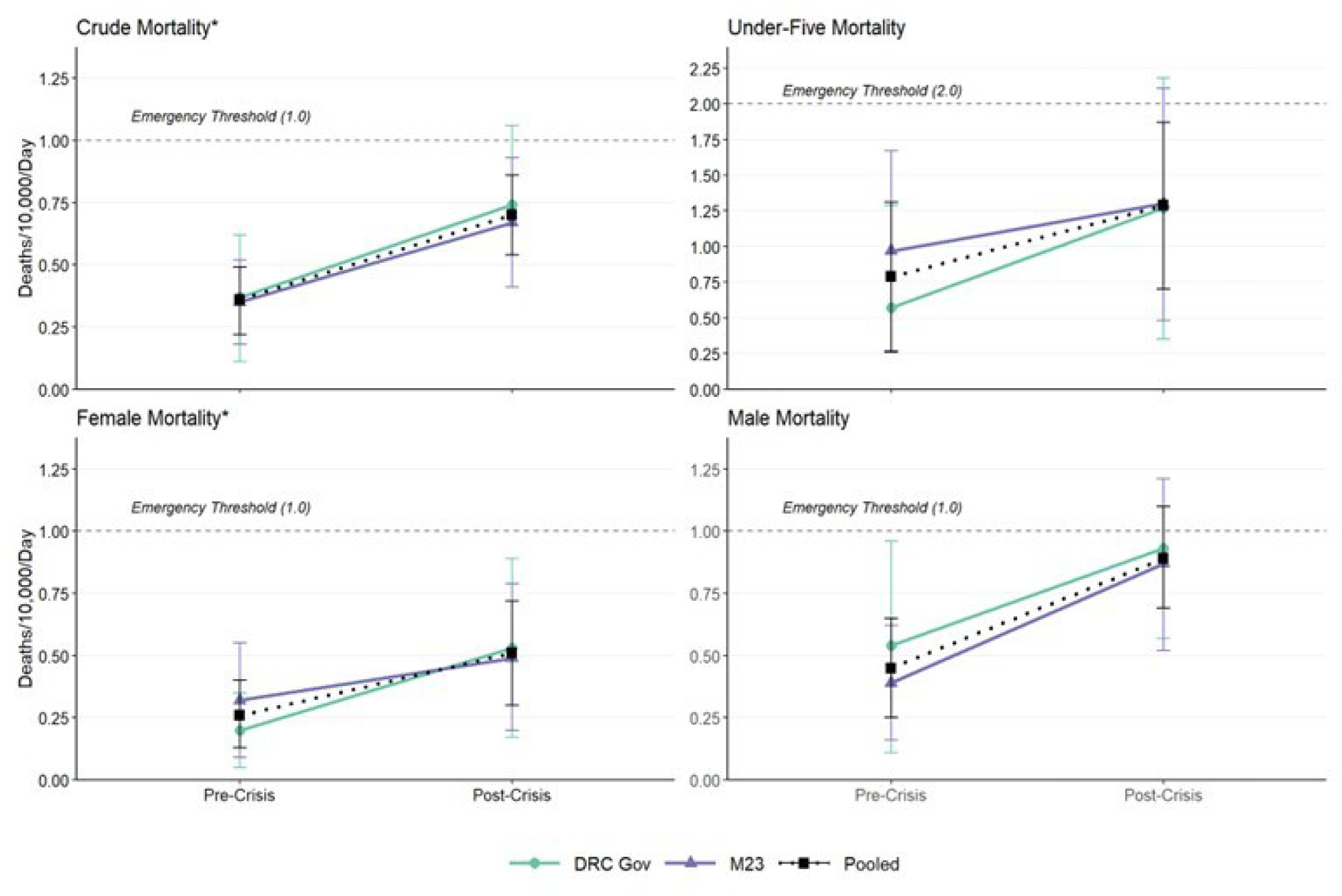
Comparison of Mortality Rates (MR) in Pre- and Post-Crisis Periods, Stratified by Territorial Control, North and South Kivu, DRC, August 2024-August 2025. Survey-weighted difference-in-differences models compared the pre-crisis baseline (1 Aug 2024–23 Jan 2025) and post-crisis period (24 Jan 2025–29 Sept 2025) for crude, under-five, female, and male mortality. Points represent marginal means adjusted for province and accessibility; vertical bars indicate 95% CIs. Asterisks (*) denote significant changes (p < 0.05). Slopes for DRC government-controlled (green circle) and M23-controlled (purple triangle) territories show uniform increases. Pooled rates (black square) increased significantly for crude (aIRR 2.02, 95% CI: 1.07, 3.80; p = 0.030) and female mortality (aIRR 2.65, 95% CI: 1.15, 6.12; p = 0.02). Non-significant DiD interaction terms for all groups indicate mortality trends were consistent regardless of territorial control.

In both unadjusted and adjusted DiD models, the transition to the post-crisis period was the only significant predictor of increased mortality. The interaction between time period and territorial control was not significant for any group (p>0.05). The results indicate the rise in mortality was geographically and temporally consistent regardless of which entity held territorial control.

Sensitivity analyses confirmed the stability of findings (Table S5). Notably, in the washout analysis excluding the month prior to and following the aid withdrawal and M23 invasion, mortality increases became more statistically robust. The time-trend reached borderline significance for male mortality (p=0.048) and approached it for children under-five (p = 0.066).

### Excess Mortality

Applying the change in crude mortality rate to the estimated population of 19.8 million, we calculated the human cost of the crisis. Analyses estimate that approximately 165,391 excess deaths (95%CI: 63,557, 267,225) occurred in North and South Kivu during the post-crisis period. If rates remain stable,this equates to 245,391 (95%CI: 94,302, 396, 492) excess deaths per year.

Direct violence accounted for 2.7% (n=1) of the 37 pre-crisis deaths and 6.6% (n=6) of the 91 post-crisis deaths. If we consider all violent deaths as excess death, we estimate that a minimum of 10,905 deaths (95% CI: 4,191, 17,619) deaths were directly attributable to violence. If we look at the change in violent deaths as a proportion of total deaths, an additional 6,435 (95%CI: 2,473, 10,397) occurred because of additional risk posed by the conflict. There was a six-fold absolute increase in violent deaths and a 2.4% increase as a proportion of all deaths.

In the analysis of proximate determinants of mortality and morbidity (Table S3), the leading household-reported causes of death were malaria/fever (n=40; 30.7% [95% CI 23.8, 38.6]), neonatal causes (n=15; 11.7% [6.7, 19.6]), and diarrhea/gastrointestinal illness (n=12; 10.0% [5.4, 17.8]). Violence was identified as the sixth most common cause of death (n=7; 5.6% [2.3, 12.7]).

The overall prevalence of global acute malnutrition (GAM) was 6.0% (n=46; 3.8, 9.3), with severe acute malnutrition (SAM) accounting for 47.8% of these cases (n=22; 2.9% [1.5, 5.3]). Disparities in malnutrition were observed by territorial control; GAM and SAM prevalence were higher in DRC-government controlled areas (n=32, 9.8% [5.6, 16.5] and n=18, 5.5% [2.8, 10.6], respectively) compared to M23-controlled areas (n=14, 3.2% [1.7, 6.0], p=0.007; and n=4, 0.9% [0.4, 2.4], p=0.006).

Additionally, 13.0% of the sample (n=754; 11.6, 14.7) reported a severe illness in the previous 30 days. Among those reporting illness, 30.6% (n=234; 24.7, 37.2) did not seek medical care, with 71.6% (n=169; 63.5, 78.4) citing financial barriers as the primary reason for forgoing treatment.

## Key Informants

Of 60 clusters visited, health officials were interviewed in 46 locations. Of the 46, three clinics had closed and only ten had received any supplies or financial support since February. Clinics reported compensating for this by raising patient user fees and laying off staff. Of the 46 informants, 27 reported changes in attendance numbers from 2024, with average weekly attendance dropping by 40% (median 35%). When treatment was sought, officials described poor quality of care due to disrupted drug supply and low staffing. Only three clinics reported no decline in attendance or quality of service. All three were still receiving international funding support.

## Discussion

The post-crisis doubling of crude mortality in the Kivus represents a systemic shock, indicating a population in acute crisis with no relief in sight. The post-crisis aIRR, the most robust measure in the analysis, provides strong evidence of the combined effects of violence, mass displacement, funding withdrawal, and health system breakdown. The rise in mortality was consistent across strata, data sources, and sensitivity analyses.

These figures almost certainly underestimate the true human cost of the crisis. Active combat rendered nine clusters inaccessible during data collection and one cluster was entirely abandoned. These clusters likely experienced the greatest burden of the crisis but remained unmeasured. Fear of reprisals would have led to underreporting of violent deaths. Conducting household interviews in communities meant we missed combatant deaths. Any households that were entirely killed, disappeared, or dissolved would also have been missed. Given the complete abandonment of one cluster, the risk of excluding such households is high. Previous studies in the DRC have documented underreporting of child and, particularly, neonatal deaths.^16^ The documentation of 11 missing persons, including 6 children, suggests additional uncounted deaths.

The withdrawal of funding was linked to increased mortality from causes that had been at least partially addressed through humanitarian programming. The most common causes of death, malaria, neonatal conditions, diarrheal disease, and tuberculosis were from treatable conditions, indicating a breakdown in access to quality care. Independent reports similarly documented deaths from preventable causes.^17^ Key informant interviews also confirmed the findings, describing severe disruptions in service availability, declining quality of care, and widespread barriers accessing treatment. Service collapse has also been reported by external actors. A 2025 assessment of 240 health structures found 80% were receiving no support and 45% had reduced staff.^18^

Armed group activity has further compounded the effects of the funding cuts. The increase in violent deaths likely reflects escalation of the conflict, as documented in UN reports^7^ and elsewhere.^19^ Movement restrictions, human rights violations, and threats by both sides of the conflict have disrupted population access to supply routes, agricultural land, and markets, effectively isolating communities^7^. The prevalence of under-five severe acute malnutrition exceeded emergency thresholds, seemingly affected by both the absence of nutrition assistance and the obstruction of livelihoods. Together, funding cuts and conflict-related access constraints have created conditions in which preventable illness, malnutrition, and death are widespread.

The geopolitical shifts suggest U.S. retreat from engagement in the region created a security vacuum. The retreat facilitated the violent M23 expansion into DRC territory, while at the same time diminishing capacity to withstand its effects. Recent analyses estimate that United States foreign assistance has saved tens of millions of lives over the past decades.^20^ In the absence of this funding, projections suggest 14 million additional preventable deaths will occur by 2030. The models largely did not account for the Bureau of Humanitarian Assistance, which comprised approximately one third of USAID’s budget and has been the primary source of funding in the Kivus.^21^

While a staged humanitarian reset could provide opportunity to reduce long-term dependency on aid and foster local governance, the current abrupt withdrawal lacks the attention and oversight needed to prevent a catastrophic loss of life. It is imperative that funding be restored to its previous levels to stop further preventable death. The reinstatement should be coupled with a shift toward the localization of aid to maximize impact, in line with the Grand Bargain target of directing 25% of humanitarian funding to local and national NGOs.^22^ Expert analyses estimate that local intermediaries can deliver results that are 32% more cost efficient than international ones.^23^ By bypassing expensive international intermediaries and channeling resources directly to national and local NGOs, the U.S. and other donor countries can provide support more effectively and efficiently. As an example, this study was completed for an estimated 60,000 USD. Restoring the full volume of aid while prioritizing local responders will ensure that a more sustainable approach does not come at the cost of the lives it is intended to protect.

## Limitations

In addition to the selection bias previously discussed, several other limitations should be considered when interpreting findings. The concurrent timing of events means it was not possible to isolate the impact of funding cuts from the M23 incursion. The short recall for both periods may have underpowered the study for sub-group analysis. Recall bias may be present if households forgot or excluded details of deaths. Finally, reliance on household reporting for cause of death rather than clinical verification introduces risk of misclassification.

## Conclusion/Recommendations

One year into the crisis in eastern DRC, reports indicate the situation has not improved.^18^ The estimates in this study suggest that nearly 250,000 excess deaths would occur annually in the Kivus if conditions remained unchanged. Given the continued hostilities and health system deterioration, mortality has likely increased since data collection occurred, amounting to a greater loss of life than captured here. If we attribute half of these excess deaths to the funding cuts and recognize that similar health system collapses have occurred in Afghanistan^24^, South Sudan^25^ and Mali^25^, previous estimates that the funding cuts will result in 14 million preventable deaths^20^ may be a substantial underestimate.

A large-scale restoration of aid is needed to prevent further escalation of mortality. The financial resources needed to avert these deaths, as it flowed in 2024, represents a very small fraction, >0.1%, of the U.S. gross domestic product per capita, and an even smaller share for the combined economies of the Development Assistance Committee countries.^26^ The current U.S. allocation of $161 million to the DRC falls far short of the $1.4 billion requested in the 2026 Humanitarian Response Plan.^27^

Assistance should prioritize resupplying and staffing primary healthcare and nutrition services. In the interim, unrestricted cash transfers can provide immediate relief to affected families. Continued mortality surveillance is necessary to monitor progress in lowering mortality. Overall, the resources required to avert these deaths, while meaningful to the communities affected, are a negligible share of global economic capacity, indicating the continued loss of life represents lack of political willpower rather than true financial constraint.

## Supporting information

DRC_Kivu_Supplement

## Data Availability

Due to the sensitive nature of the study, data are not publicly shared. Requests for data may be submitted to the corresponding author and considered on a case-by-case basis.

## Acknowledgements

This study was made possible through the dedication and courage of the local and national actors who designed, coordinated, and carried out data collection under exceptionally difficult conditions. The Congolese individuals who contributed their time and energy to this work did so at considerable personal risk. Several qualified for authorship but could not disclose their identities without endangering themselves and their families. Their commitment to documenting this crisis is not only a reflection of its severity, but of the determination of the people closest to it to improve the conditions in which their communities live and die.

