## Supplementary material for "Mortality in Eastern Democratic Republic of the Congo: A Population-Based Survey Following the 2025 M23 Offensive and Humanitarian Funding Withdrawal": DRC_Kivu_Supplement

#### *1. Methodological Note*

**Model Selection**

For all mortality outcomes, we compared Poisson, negative binomial (NB), and zero-inflated negative binomial (ZINB) models on cluster-level death counts with a person-time offset. Based on the Akaike Information Criterion (AIC) hypothesis tests, the Poisson distribution was found to provide the most parsimonious fit over the NB model for crude mortality as well as under-five, female, and male mortality subsets.

As an additional check, we formally tested for zero-inflation using the Vuong non-nested hypothesis test. The AIC-corrected Vuong test significantly favored the standard count model over the zero-inflated alternative, indicating the observed zeros were adequately handled by the primary model's structure and survey-design adjustments. Model selection was guided by the principle of parsimony. For the crude and male mortality models, the AIC-corrected Vuong test significantly favored the Negative Binomial over the ZINB specification ($p<0{\cdot}05$). For the under-five and female subsets, the models were statistically indistinguishable ($p >0{\cdot}05$). Given these results, along with the numerical instability of complex specifications in the male mortality subset, the Poisson distribution with robust standard errors was selected as the final model for all outcomes.

Since the standard Poisson models exhibited overdispersion, we utilized survey-weighted Poisson regression (via Taylor-series linearization) as the most stable estimation method. This approach employs a robust sandwich estimator to ensure that 95% confidence intervals account for cluster-level overdispersion. We report the dispersion parameter $\phi$, where values exceeding 1.0 indicate overdispersion. Sensitivity analyses confirmed that this specification yielded results equivalent to a Quasipoisson model, demonstrating that the robust standard errors adequately addressed the observed dispersion. Further details on model selection and diagnostic tests are provided in Table S1.

**Diagnostic Testing**

We evaluated three metrics of survey integrity to assess reliability of point estimates and precision of confidence intervals:

1. **Standard Error Specification:** Given the high dispersion parameters, robust standard errors were utilized as a conservative measure to account for non-spatial overdispersion (heteroscedasticity) that standard survey errors might underestimate. This testing was used to confirm the 95% confidence intervals were robust against divergence from model assumptions, (specifically the presence of higher-than-expected variability [overdispersion]) which could lead to misleadingly precise results.
2. **Weight Stability:** We assessed the variability of sampling weights using the Coefficient of Variation (CV). Across all models, the CV remained low (range of 0.12 to 0.13), indicating that the weights

were relatively consistent and uniform across clusters and the variance was not artificially inflated by extreme weight values.

3. **Clustering and ICC:** We calculated the Intraclass Correlation Coefficient (ICC) and the Design Effect (DEFF) at the cluster level. The ICC was estimated at 0.04 for crude mortality and 0.08 for under-five mortality meaning that 4% and 8% of the variability was explained by clustering. The DEFF remained near ~1.0 for all outcomes. However, to maintain a conservative and consistent approach we used cluster-robust estimators via Taylor-series linearization to fully account for the complex sampling structure.

**Table S1: Model Selection and Diagnostic Statistics by Mortality Outcome Group**

| | **Crude** | **Under-Five** | **Female** | **Male** |
|---|---|---|---|---|
| Model Specification | Poisson | Poisson | Poisson | Poisson |
| AIC (Poisson vs NB)* | 327·39 vs 326·94 | 166·95 vs 168·26 | 205·41 vs 205·34 | 246·11 vs 248·11 |
| AIC-Corrected Vuong Test p-value (NB vs ZINB) ǂ | <0.02 | 0.08 | 0.31 | <0.001 |
| Dispersion Basis[1] | ϕ = 14·66 | ϕ = 23·40 | ϕ = 15.12 | ϕ = 14.51 |
| Robust SE (Interaction Term)[2] | 0.36 | 0.50 | 0.35 | 0.61 |
| Design Effect[3,4] | 0.90 | 1.01 | 0.84 | 0.93 |
| Adjusted ICC[3] | 0.04 | 0.08 | 0.06 | 0.01 |
| CV Weights[5] | 0.13 | 0.12 | 0.12 | 0.13 |

**AIC** = Akaike Information Criterion; **NB** = Negative Binomial; **ZINB** = Zero-Inflated Negative Binomial; **SE** = Standard Error; **ICC** = Intraclass Correlation Coefficient; **CV** = Coefficient of Variation

[*] AIC (Poisson vs. NB): Akaike Information Criterion; lower values indicate a more parsimonious fit. In all cases, Poisson was favored.

[ǂ]*AIC-Corrected Vuong: Compares the Negative Binomial (NB) to the Zero-Inflated (ZINB). A p < 0.05 (Crude, Under-Five, Male) indicates NB is superior; p>0.05 (Female) indicates the models are indistinguishable, favoring parsimony.*

[1]*Dispersion Basis: Values represent the dispersion parameter ϕ; values > 1.0 indicates overdispersion.*

[2]*Robust SE: Non-exponentiated standard error of the interaction term, calculated via Taylor-series linearization using the sandwich estimator from svyglm to ensure conservative 95% CIs in the presence of overdispersion.*

[3]*Design Effect and ICC were calculated at the village (cluster) level representing proportion of total variance attributed to cluster*

[4]*Design effect calculated for the unadjusted model to demonstrate the impact of cluster sampling on variance.*

[5]*CV Weights: Values <1.0 suggest weighting did not excessively inflate variance*

### *2. Difference-in-Differences Poisson Distribution Model Population Mortality*

**Table S2.** Difference-in-Differences Poisson Distribution Model Population Mortality
*Key Finding: Described in main article.*

| | **aIRR** | **95%CI:** | **p-value** |
|---|---|---|---|
| **Crude** | | | |
| Post-crisis Period | 2.02 | 1.07, 3.80 | 0.03 |
| M23-Controlled Area | 0.96 | 0.37, 2.48 | 0.93 |
| Province: North Kivu | 0.95 | 0.56, 1.61 | 0.84 |
| Accessibility: Hard-to-Reach | 1.42 | 0.70, 2.86 | 0.33 |
| Interaction: Post-crisis × M23-Controlled Area | 0.95 | 0.46, 1.96 | 0.88 |
| **Under-Five** | | | |

| | | | |
|---|---|---|---|
| Post-crisis Period | 2.23 | 0.55, 9.10 | 0.26 |
| M23-Controlled Area | 1.71 | 0.39, 7.55 | 0.47 |
| Province: North Kivu | 0.44 | 0.15, 1.25 | 0.12 |
| Accessibility: Hard-to-Reach | 0.66 | 0.28, 1.55 | 0.33 |
| Interaction: Post-crisis × M23-Controlled Area | 0.60 | 0.13, 2.80 | 0.51 |
| **Female** | | | |
| Post-crisis Period | 2.65 | 1.15, 6.12 | 0.023 |
| M23-Controlled Area | 1.58 | 0.53, 4.68 | 0.40 |
| Province: North Kivu | 0.95 | 0.46, 1.99 | 0.90 |
| Accessibility: Hard-to-Reach | 1.91 | 0.79, 4.65 | 0.15 |
| Interaction: Post-crisis × M23-Controlled Area | 0.59 | 0.19, 1.85 | 0.36 |
| **Male** | | | |
| Post-crisis Period | 1.74 | 0.82, 3.68 | 0.15 |
| M23-Controlled Area | 0.73 | 0.25, 2.09 | 0.55 |
| Province: North Kivu | 0.94 | 0.56, 1.59 | 0.82 |
| Accessibility: Hard-to-Reach | 1.18 | 0.60, 2.30 | 0.63 |
| Interaction: Post-crisis × M23-Controlled Area | 1.27 | 0.50, 3.27 | 0.61 |

**aIRR =** Adjusted Incident Rate Ratio; **95%CI:** = Confidence Interval
**Statistical Analysis:** Estimates were derived from survey-weighted Poisson regression models adjusted for province and accessibility. The analysis incorporated inverse probability weights and cluster-robust standard errors to account for the sampling design and intra-cluster correlation at the village and household levels.
**Significance:** Statistical significance is set at p<0.05

### *3. Mortality Rates by Territorial Control*

**Table S3. Adjusted Mortality Rates Stratified by Territorial Control and Crisis Period in North and South Kivu, DRC, August 2024- August 2025.**

*Key Finding: Crude mortality increased during the post-crisis period, regardless of territorial governance. While the aggregate study population showed a significant increase in mortality (as reported in the main text), the pre-to-post change within individual territorial strata and within population subgroups did not reach statistical significance due to the reduced power of the subdivided samples. However, the high degree of similarity in change in DRC-Government-controlled and M23-controlled areas as confirmed by a non-significant Difference-in-Differences interaction in all groups (p>0.05) indicates that the change was consistent across both zones.*

| | DRC-Government Controlled | | M23-AFC Controlled | | |
|---|---|---|---|---|---|
| | **Pre-Crisis Rate (95%CI)** | **Post-Crisis Rate (95%CI)** | **Pre-Crisis Rate (95%CI)** | **Post-Crisis Rate (95%CI)** | **Post-Crisis × M23 p-value** |
| Crude | 0.38 (0.12, 0.65) | 0.78 (0.49, 1.06) | 0.37 (0.16, 0.58) | 0.71 (0.38, 1.04) | 0.88 |
| Under-Five | 0.51 (0.00, 1.15) | 1.14 (0.36, 1.92) | 0.87 (0.18, 1.56) | 1.16 (0.33, 2.00) | 0.51 |
| Female | 0.22 (0.06, 0.39) | 0.59 (0.24, 0.93) | 0.35 (0.08, 0.62) | 0.559 (0.19, 0.90) | 0.36 |
| Male | 0.55 (0.13, 0.97) | 0.96 (0.64, 1.27) | 0.40 (0.15, 0.65) | 0.88 (0.47, 1.30) | 0.61 |

**95%CI:** = Confidence Interval; **DiD** = Difference- in-Differences
**Rate Units:** Adjusted mortality rates are expressed as deaths per 10,000 person-days.
**Statistical Analysis:** Estimates were derived from survey-weighted Poisson regression models adjusted for province and accessibility. The analysis incorporated inverse probability weights and cluster-robust standard errors to account for the

sampling design and intra-cluster correlation at the village and household levels. Mortality rates represent adjusted marginal mean rates (deaths/10,000/day) from the survey-weighted Poisson models.

**Significance:** Statistical significance is set at $p<0.05$

**Clustering:** Statistical analyses incorporated inverse probability weights to account for sampling design and cluster-robust standard errors to account for the intra-cluster correlation at the village and household levels.

### *4. Sensitivity Analyses*

**Table S4: Model Stability and Sensitivity Analysis of Difference-in-Differences Mortality Risk Estimates in North and South Kivu, DRC, August 2024-August 2025**

*Key Findings: Robustness of Crude and Female Time-Trends: The overall mortality increase (Time-Trend) remained consistently significant across sensitivity testing for crude mortality (p= 0.030) and female mortality (p=0.020). The sensitivity analyses reinforce the primary findings and confirm stability of the results regardless of model specifications.*

*Consistency of Estimates: The point estimates (aIRRs) showed strong stability with design effects remaining near 1.0 for all sub-groups across sensitivity testing.*

*Robustness of aIRR: Across all sensitivity testing, the Interaction aIRR remained non-significant (p>0.05), indicating the lack of divergence in mortality between territorial control zones is a stable finding, unaffected by the inclusion of geographic covariates or high-influence death clusters.*

*Impact of the Washout Period (Jan/Feb Exclusion): While the time-trend for males was non-significant in the primary model (p = 0.15), it reached borderline statistical significance when deaths ±1 month from the crisis were removed (p = 0.048). This suggests that short-term volatility in the transition period may have masked an upward mortality trend for males. Similarly, the time-trend for children under five moved from a primary value of p = 0.26 to p = 0.066 (just above the significance threshold). The finding indicates that the mortality increase in children became more pronounced and consistent when deaths around the acute crisis period were excluded.*

| | Post-Crisis × M23 aIRR (95%CI) | Post-Crisis × M23 p-value | Post-Crisis Period aIRR (95%CI) | Post-Crisis Period p-value | Design effect |
|---|---|---|---|---|---|
| **Crude** | | | | | |
| Primary Adjusted Model | 0.95 (0.46, 1.96) | 0.88 | 2.02 (1.07, 3.80) | 0.030 | 0.90 |
| Unadjusted Model | 0.94 (0.46, 1.95) | 0.87 | 2.04 (1.08, 3.83) | 0.028 | 0.90 |
| Exclude Access Covariate | 0.95 (0.46, 1.96) | 0.88 | 2.03 (1.08, 3.83) | 0.029 | 0.91 |
| Exclude Highest Death Cluster(s) | 0.84 (0.37, 1.93) | 0.68 | 2.39 (1.16, 4.96) | 0.020 | 0.98 |
| Exclude Deaths ± 1 month of Crisis Date | 0.72 (0.32, 1.63) | 0.42 | 2.64 (1.31, 5.36) | 0.008 | 0.91 |
| **Under-Five** | | | | | |
| Primary Adjusted Model | 0.60 (0.13, 2.80) | 0.51 | 2.23 (0.55, 9.10) | 0.26 | 1.01 |
| Unadjusted Model | 0.58 (0.13, 2.69) | 0.48 | 2.26 (0.56, 9.09) | 0.25 | 1.01 |
| Exclude Access Covariate | 0.60 (0.13, 2.78) | 0.51 | 2.21 (0.55, 8.94) | 0.26 | 1.00 |
| Exclude Highest Death Cluster(s) | 0.63 (0.13, 3.14) | 0.57 | 2.21 (0.54, 8.98) | 0.26 | 1.03 |
| Exclude Deaths ± 1 month of Crisis Date | 0.28 (0.03, 2.28) | 0.23 | 6.26 (0.88, 44.4) | 0.07 | 0.85 |
| **Female** | | | | | |
| Primary Adjusted Model | 0.59 (0.19, 1.85) | 0.36 | 2.65 (1.15, 6.12) | 0.02 | 0.84 |
| Unadjusted Model | 0.59 (0.18, 1.85) | 0.36 | 2.68 (1.16, 6.18) | 0.02 | 0.85 |

| | | | | | |
|---|---|---|---|---|---|
| Exclude Access Covariate | 0.59 (0.19, 1.86) | 0.36 | 2.67 (0.022) | 0.02 | 0.85 |
| Exclude Highest Death Cluster(s) | 0.63 (0.17, 2.26) | 0.47 | 2.64 (1.14, 6.11) | 0.02 | 0.85 |
| Exclude Deaths ± 1 month of Crisis Date | 0.54 (0.15, 2.00) | 0.35 | 3.06 (1.14, 8.22) | 0.03 | 0.93 |
| **Male** | | | | | |
| Primary Adjusted Model | 1.27 (0.50, 3.27) | 0.61 | 1.74 (0.82, 3.68) | 0.15 | 0.93 |
| Unadjusted Model | 1.27 (0.50, 3.25) | 0.61 | 1.75 (0.83, 3.69) | 0.14 | 0.93 |
| Exclude Access Covariate | 1.27 (0.50, 3.27) | 0.61 | 1.74 (0.82, 3.69) | 0.61 | 0.93 |
| Exclude Highest Death Cluster(s) | 1.03 (0.36, 2.92) | 0.96 | 2.15 (0.90, 5.16) | 0.08 | 1.01 |
| Exclude Deaths ± 1 month of Crisis Date | 0.88 (0.28, 2.788) | 0.83 | 2.42 (1.01, 5.84) | 0.05 | 1.05 |

**aIRR=**Adjusted Incidence Rate Ratio; **95%CI:** = Confidence Interval; **DiD**= *Difference-in-Differences*

**Statistical Analysis:** Estimates were derived from survey-weighted Poisson regression models adjusted for province and accessibility. The analysis incorporated inverse probability weights and cluster-robust standard errors to account for the sampling design and intra-cluster correlation at the village and household levels. The aIRR reflects the regional mortality change between pre-crisis (Aug 2024–Jan 2025) and post-crisis (Feb–Aug 2025) periods. The interaction p-value tests whether changes in mortality differed significantly between DRC Government and M23-AFC territorial control.

**Significance:** Statistical significance is set at $p<0.05$

**Design Effect:** Calculated for the unadjusted model to demonstrate the impact of cluster sampling on variance.

### *5. Additional Descriptive Data*

#### Age Pyramids

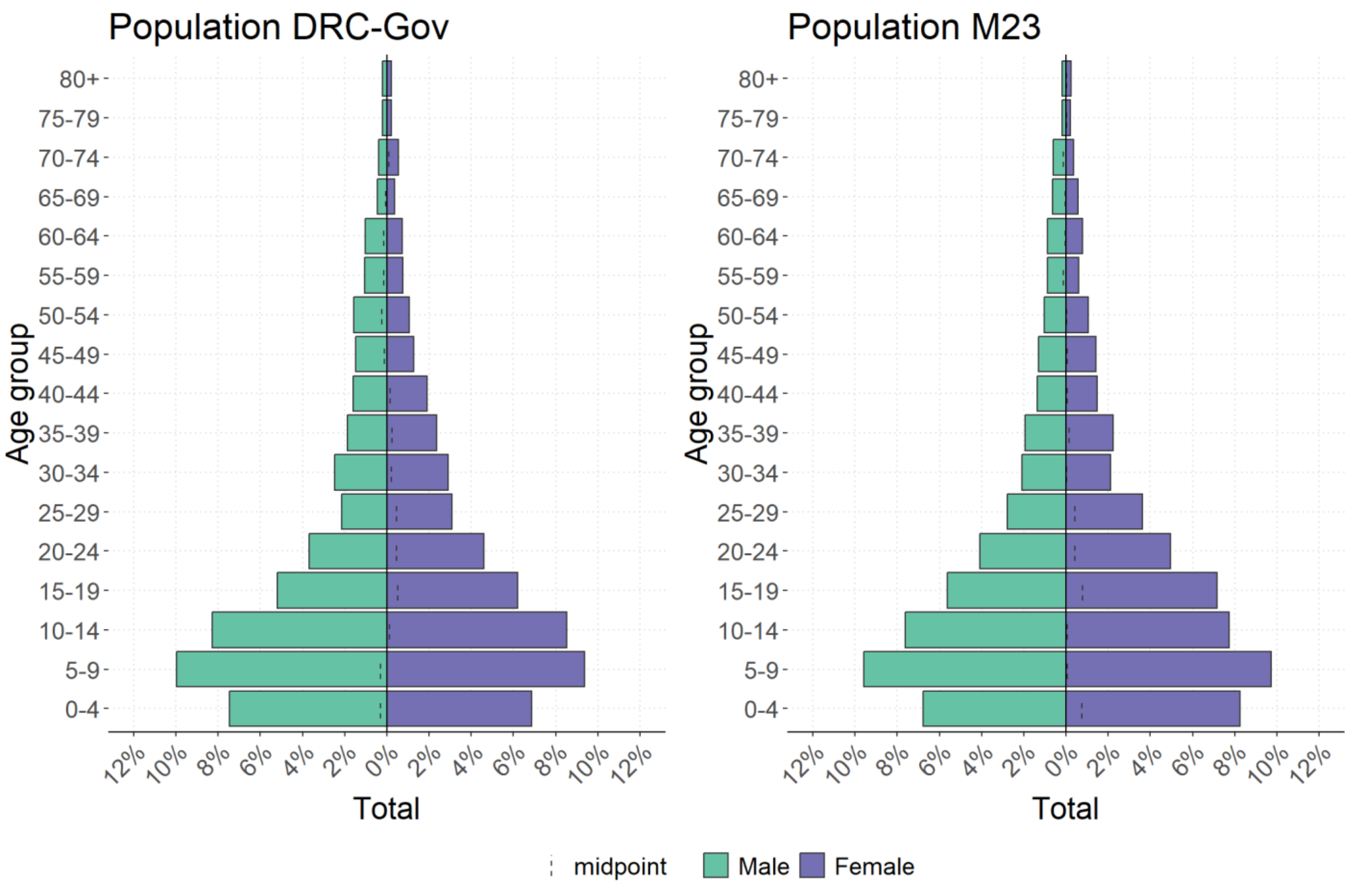


***Figure 1. Population Structure of the Surveyed Households.*** *The age-sex pyramid displays the demographic distribution of the study population, with males represented in green (left) and females in purple (right).*

### Birth Rates

**Table S5. Crude Birth Rate, Stratified by Territorial Control and Crisis Period in North and South Kivu, DRC, August 2024–August 2025**

*Key Finding: While no significant differences in crude birth rates were observed between DRC-Government and M23 controlled territory, both areas experienced a statistically significant increase in birth rates during the post-crisis period (p < 0.001). Overall, the birth rate rose from 10.7 per 1,000 (95% CI 7.8, 13.6) pre-crisis to 19.7 per 1,000 (95% CI 16.1, 23.2) post-crisis.*

| | n | Pre-Crisis Rate (95%CI) | n | Post-Crisis Rate (95%CI) | p-value |
|---|---|---|---|---|---|
| Overall | 73 | 10·7 (7·8, 13·6) | 135 | 19·7 (16·1, 23·2) | <0.001 |
| DRC-Gov | 28 | 10·2 (6·0, 14·5) | 44 | 16·0 (10·3, 21·7) | <0.001 |
| M23 | 45 | 11·1 (7·2, 15·0) | 91 | 22·1 (17·7, 26·5) | <0.001 |

**95%CI:** = Confidence Interval

**Rate Units**: Birth rates expressed as births per 1,000 population

**Model Specification:** Estimates were derived from survey-weighted Poisson regression models (svyglm) incorporating household-level weights.

### *5. Secondary Outcomes*

**Table S6. Proximate Determinants of Mortality, Morbidity and Health Service Utilization by Territorial Control**

*Key Findings: While malaria remained the leading cause of death across both regions, M23 controlled areas reported significantly more deaths from neonatal causes. Other differences in cause of death remained non-significant between territories. Significant health disparities were found between territories. Global Acute Malnutrition (GAM) and Severe Acute Malnutrition (SAM) were significantly more prevalent in DRC-Government areas (9·8% and 5·5%, respectively) than in M23-controlled zones (3·2%, p=0·007; and 0·9%, p=0·006). Despite varying disease burdens, health-seeking behavior was consistent across the sample, with nearly one-third of ill individuals not seeking care for severe illness (30·6%), primarily due to prohibitive financial barriers (71·6%).*

| | Total | | DRC – Gov | | M23 – AFC | |
|---|---|---|---|---|---|---|
| | ***n*** | **% (95%CI)** | ***n*** | **% (95%CI)** | ***n*** | **% (95%CI)** |
| **Household Reported Cause of Death** | | | | | | |
| Malaria/Fever | 40 | 30·7 (23·8, 38·6) | 20 | 31·7 (21·7, 43·8) | 20 | 29·7 (20·8, 40·5) |
| Neonatal | 15 | 11·7 (6·7, 19·6) | 2 | 3·1 (0·9, 10·0) | 13 | 20·0 (11·1, 33·3) |
| Diarrhea/Gastrointestinal | 12 | 10·0 (5·4, 17·8) | 8 | 14·0 (6·7, 26·9) | 4 | 6·2 (2·3, 15·5) |
| Hypertension/Cardiac | 8 | 6·3 (3·3, 11·9) | 4 | 6·7 (2·6, 16·2) | 4 | 6·0 (2·4, 14·4) |
| Diabetes | 7 | 5·7 (2·7, 11·4) | 6 | 10·0 (4·6, 20·4) | 1 | 1·5 (0·2, 9·7) |
| Violence | 7 | 5·6 (2·3, 12·7) | 3 | 4·9 (1·4, 15·6) | 4 | 6·2 (1·8, 18·9) |
| Trauma/Accident | 5 | 4·0 (1·7, 9·3) | 1 | 1·6 (0·3, 9·6) | 4 | 6·3 (2·3, 16·4) |
| Tuberculosis | 5 | 3·7 (1·3, 10·1) | 5 | 7·6 (2·8, 19·2) | 0 | 0·0 (0·0, 0·0) |
| Respiratory Infection | 4 | 3·0 (0·9, 9·5) | 4 | 6·2 (2·0, 17·6) | 0 | 0·0 (0·0, 0·0) |
| Sudden Death | 4 | 3·0 (1·1, 7·8) | 1 | 1·5 (0·2, 9·7) | 3 | 4·5 (1·4, 13·3) |
| Other Causes | 21 | 16·2 (9·5, 26·3) | 8 | 12·7 (5·2, 27·6) | 13 | 19·6 (10·1, 34·5) |
| **Malnutrition** | | | | | | |
| Prevalence of Global Acute Malnutrition | 46 | 6·0 (3·8, 9·3) | 32 | 9·8 (5·6, 16·5) | 14 | 3·2 (1·7, 6·0) |
| Prevalence of Severe Acute Malnutrition | 22 | 2·9 (1·5, 5·3) | 18 | 5·5 (2·8, 10·6) | 4 | 0·9 (0·4, 2·4) |
| **Health-Seeking Behavior** | | | | | | |
| Reported illness in past 30 days | 754 | 13·0 (11·6, 14·7) | 354 | 15·1 (12·9, 17·6) | 400 | 11·7 (9·8, 13·6) |
| Did not seek care for illness | 234 | 30·6 (24·7, 37·2) | 121 | 33·5 (24·7, 43·7) | 113 | 27·8 (20·6, 36·4) |
| Cited cost as the main barrier to care | 169 | 71·6 (63.5, 78.4) | 92 | 74.7 (66.3, 81.6) | 77 | 68.0 (53.9, 79.5) |
